# Comparison of Self-Collected Vaginal Swabs and First-Void Urine for Detection of Human Papillomavirus in Sexually Active Girls and Women in Three South Asian Countries

**DOI:** 10.1101/2025.08.01.25332560

**Authors:** Nigus Telele, Nishan Katuwal, Furqan Kabir, Sezanur Rahman, Sabin Bikram Shahi, Raja Ram Yadav, Fatima Aziz, Md Abir Hossain, Alexandra M. Hill, Indriana Permatasari, Yubin Lee, Haeun Cho, Jiyoung Lee, Young Ae You, Hyeon Seon Ahn, Soim Park, Dipesh Tamrakar, Aneela Pasha, Anadil Alam, Muslima Ejaz, M Imran Nisar, Troy D. Querec, Bolarinde J. Lawal, Edward M. Choi, Deborah Watson-Jones, Pritha Basnet, Rajeev Shrestha, Abha Shrestha, Fyezah Jehan, Quamrun Nahar, Aneeta Hotwani, Mustafizur Rahman, Julia A. Lynch, Elizabeth R. Unger

## Abstract

**Background:** As more countries plan and launch human papillomavirus (HPV) vaccination campaigns, reliable self-sampling methods are essential for detecting high-risk HPV (HR-HPV) types and assessing public health impact. This study compared HR-HPV detection in self-collected vaginal swabs (SCVS) and first-void urine (FVU) among sexually active girls and women aged 15-25 years in Bangladesh, Nepal, and Pakistan.

**Methods:** Within the Global Burden Estimation of HPV (GLOBE-HPV) project, which aims to estimate HPV prevalence and incidence across eight low- and middle-income countries in South Asia and Sub-Saharan Africa, we analyzed paired SCVS and FVU samples from 753 participants in Bangladesh, Pakistan, and Nepal using standardized protocols. DNA was extracted using the QIAamp DNA Mini Kit, and HPV testing was performed with the Allplex HPV28 Detection PCR assay. We evaluated HPV detection and type-specific concordance using Cohen’s Kappa, McNemar’s test, and a 3×3 agreement table, along with accuracy and positive/negative agreement metrics.

**Results:** The overall prevalence of 14 HR-HPV types was 8.6% in SCVS and 7.2% in FVU samples. Detection rates for 7 HR-HPV vaccine types were similar (5.3% in SCVS versus 5.0% in FVU), with nearly identical HPV 16/18 rates (2.3% in SCVS and 2.4% in FVU). Bi-directional type-specific discordance was noted, with each sample type detecting unique types. SCVS demonstrated higher sensitivity for detecting HPV types beyond the 9 vaccine types with McNemar p-values of 0.013 and <0.001 for 14 and 28 types, respectively, while overall concordance remained high (Kappa >0.7). Samples with lower viral load (indicated by higher real-time PCR cycle threshold values) were more likely to yield discordant results.

**Conclusion:** SCVS and FVU yielded similar HR-HPV results, including those targeted by the 9-valent HPV vaccine, in sexually active young women. Both non-invasive self-sampling methods have potential for use in large-scale HPV surveillance programs in resource-limited settings.

## Introduction

Human papillomavirus (HPV) is the most common sexually transmitted infection worldwide, with approximately 85% of sexually active individuals acquiring it during their lifetime (1). This double-stranded DNA virus, belonging to the *Papillomaviridae* family, is classified into high-risk (HR-HPV) and low-risk (LR-HPV) types based on oncogenic potential (2). While most HPV infections clear naturally within two years, persistent HR-HPV infections—specifically those caused by the 14 oncogenic types, including HPV 16 and 18, which account for roughly 70% of cervical cancers—can lead to cervical, vulvar, and anal cancers (3–5). In contrast, low-risk types such as HPV 6 and 11 are mainly associated with benign conditions like genital warts. Recognizing HPV as the primary cause of cervical cancer has spurred critical advancements in screening and vaccine development, significantly contributing to global prevention efforts (6).

Cervical cancer is the fourth most common cancer worldwide, with a disproportionate burden in low- and middle-income countries (LMICs), particularly in Africa and Asia (7). There are highly effective vaccines to prevent infection and high rates of cure with early detection. However, South Asian countries, including Bangladesh, Pakistan, and Nepal, face significant challenges in reducing cervical cancer burden due to limited healthcare infrastructure, insufficient data on HPV burden, low rates of HPV screening, and limited access to vaccine (8, 9).

In Bangladesh, cervical cancer is the second most common cancer among women, with 9,640 new cases and 5,826 deaths reported in 2022 (10, 11). A 2014 population-based study reported an overall HPV prevalence of 7.7% among ever-married women aged 13–64 years, with no significant difference between urban (7.9%) and rural (7.4%) participants. The most common HR-HPV types identified were HPV16, HPV66, HPV18, HPV45, HPV31, and HPV53 (12). Despite a screening program for women aged 30–60 years, coverage remains low, and it mainly uses visual inspection with acetic acid (VIA) instead of HPV DNA testing (8, 13). Bangladesh introduced HPV vaccination targeting girls 10-14 years of age in October 2023 (14).

Similarly, cervical cancer is the second most common cancer among women under 50 in Pakistan and the third leading cause of cancer-related deaths in reproductive-aged women. Over 70% of cancer patients report with very advanced stage of malignancy. Pakistan has not yet introduced the HPV vaccine (15, 16).

In Nepal, cervical cancer is the leading cancer among women, with an incidence of 16.4 per 100,000 and a high mortality rate, driven by HPV types 16 and 18 (17, 18). Despite efforts using VIA, only 8.2% of women aged 30-49 were screened by 2019, far below the 70% target (19). In February 2025, Nepal introduced HPV vaccines into their national program targeting girls 10-15 years of age (20).

Due to the slow progression from infection to development of cervical cancer, it may take decades to measure the impact of HPV vaccination programs on cancer reduction. Therefore, sampling tools that enable HPV DNA detection are valuable for monitoring program effectiveness over shorter periods. Self-collection methods, such as vaginal swabs and first-void urine (FVU) samples, are gaining attention as alternatives to healthcare providers-collected samples and are suitable for population-based surveillance (21, 22). Studies have shown that self-collected samples can reliably detect HR-HPV types, offering a more accessible and less invasive option (23–26). They are especially valuable for their ease of use, privacy, and ability to reach underserved populations (27).

Self-collected vaginal swabs (SCVS) show high concordance with clinician-collected cervical swab samples for HPV detection (28). For example, in a study of 245 women referred to colposcopy for abnormal cytology, with likely higher viral loads than in screening population, hr-HPV detection concordance between clinician and self-collected vaginal samples was high (k=0.898) (29). Another study done by Qi *et al*. involving volunteer women aged ≥18 years who self-collected vaginal swabs for HPV testing under various conditions demonstrated that SCVS were highly concordant with clinician-collected cervical samples for HPV detection, with a total agreement of 90.3% and a positive percentage agreement of 84.2% (30).

First-void urine, with its ease of collection and non-invasive nature is another option for self-sampling (21, 31–36). In some cultures, FVU sampling may be preferred over cervical or vaginal sampling (21, 22, 35). A systematic review and meta-analysis found that compared to cervical samples, FVU has a pooled sensitivity of 78% and specificity of 89% for detecting HR-HPV and particularly high sensitivity for HPV16 (77%) and HPV18 (98%) (25). Devices like Colli-Pee® help standardize FVU collection, ensuring consistent and reliable results (29, 35, 37). However, more research is needed to verify that urine-based self-sampling is suitable for population-based surveillance. Therefore, this study aimed to assess and compare the effectiveness of SCVS and FVU for detecting HR-HPV from sexually active girls and women aged 15-25 years in three South Asian countries.

## Materials and Methods

### Study design

The Global Burden Estimation of HPV (GLOBE-HPV) study is a multi-country, multi-site project aimed at estimating the burden of HPV among girls and women in 8 GAVI-eligible countries in South Asia (Bangladesh, Pakistan, and Nepal) and sub-Saharan Africa (Democratic Republic of Congo, Ghana, Sierra Leone, Tanzania, and Zambia). One of its particular aims is to assess the incidence of persistent HPV infection through longitudinal cohort studies among sexually active girls and young women (38). During the baseline visit of the longitudinal cohorts, paired SCVS and FVU samples were collected to evaluate the concordance of the resulting HPV type detection. This paper reports the results of HPV prevalence and concordance between the two self-sampling methods from the three South Asian countries. This study is registered on clinicaltrials.gov (39).

### Study setting and period

This concordance study was conducted in low-income urban and peri-urban communities across Bangladesh, Pakistan, and Nepal. Sample collection and testing were completed from November 2023 to September 2024. The participating institutions, study sites, and study population are summarized in Table 1.

**Table 1.**
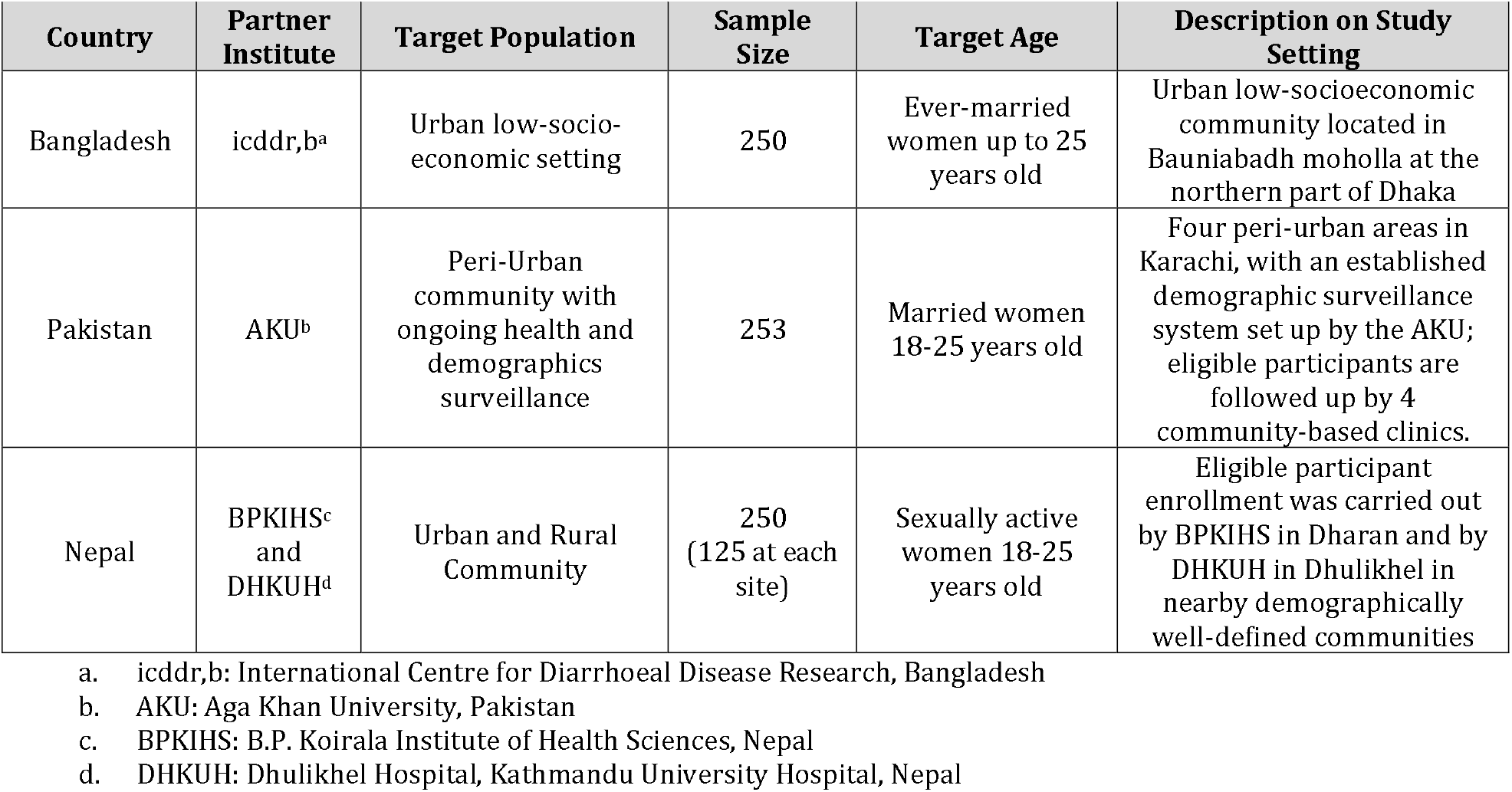
Study sites, study population, and sample size by country.

In Bangladesh, the study was conducted in Bauniabadh, Mirpur, Dhaka, a densely populated (50,000 people/km^2^), low-socioeconomic community with limited healthcare access. The study enrolled 250 ever-married women (≤25 years old) through stratified systematic sampling. In Pakistan, 253 participants were recruited from four peri-urban coastal slum communities in Karachi including Rehri Goth, Bhains Colony, Ibrahim Hyderi, and Ali Akber Shah, which have a combined population of approximately 350,000. Enrollment was based on established line listings from a demographic surveillance system (DSS). In Nepal, 250 participants were selected from urban and rural catchment areas of BPKIHS and DHKUH, with recruitment conducted in well-baby/immunization clinics and through community networks using purposive sampling.

### Inclusion and exclusion criteria

Girls and women were deemed eligible if they met the following criteria: aged within the target range, sexually active, residents of the study community for at least the past 3 months, and able to understand the purpose of the study and study procedures. When eligible for the longitudinal cohort girls/women provided informed consent or assent to provide samples every 6 months over the course of 2 years. In Bangladesh, participants were required to be currently or previously married and aged 25 years or younger. In Pakistan, participants were required to be married and aged 18 to 25 years due to cultural and legal reasons. In Nepal, women aged 18-25 years were eligible if they were sexually active, regardless of marital status. The exclusion criteria included refusal to participate in any aspect of the study or the presence of a medical condition or other factor that, in the opinion of the investigator, precluded study participation.

### Sample collection and test procedure

This sub-study was conducted under a harmonized sampling and laboratory protocol to ensure comparability of results from participating countries. Data collectors trained participants in self-collection of SCVS and FVU samples in the local languages through detailed verbal explanations, diagrams, and demonstrations and aided, if requested, to ensure proper placement of devices. The SCVS samples were collected using the Aptima® Multitest Swab Specimen Collection Kit. The FVU samples were collected following the manufacturer’s instructions using a 10-mL Colli-Pee® device with 3 mL of urine conservation medium (UCM). First-void urine samples could be collected at any time of the day. Participants who had urinated within the previous hour were instructed to wait one hour before collection. For women who were menstruating on the day of their scheduled visit, the sample collection was typically rescheduled to avoid any potential effect of blood in HPV testing. A total of 753 paired SCVS and FVU samples were collected, 250 from Bangladesh, 253 from Pakistan, and 250 from Nepal (Table 1).

Once collection was completed, samples were stored in cool boxes (2–8°C) and transported to the central laboratory in each country with continuous temperature monitoring to ensure stability. At the respective central laboratories (icddr,b in Bangladesh, IDRL-AKU in Pakistan, and DHKUH in Nepal), samples were accessioned and checked for quality, noting any abnormalities (e.g. missing samples, reduced volume, blood, leakage, etc.). SCVS samples were stored at −20°C. FVU samples were centrifuged at 500 rcf for 5 minutes, 9 mL removed, pellet resuspended in residual one mL of supernatant and divided into two 0.5-mL aliquots. The aliquot for DNA extraction was stored at −20°C, and the one for long-term storage was kept at −80°C. In Nepal, to avoid degradation from delays in processing during long distance transportation to the central testing laboratory, samples collected at BPKIHS were accessioned and processed locally.

DNA extraction from both sample types was performed manually using the QIAamp® DNA Mini Kit following the manufacturer’s instructions, with quality controls including water blanks in each extraction run. The AllPlex™ 28 HPV Detection PCR assay on a Bio-Rad CFX96 Dx system, detecting 28 HPV types (HPV 6, 11, 16, 18, 26, 31, 33, 35, 39, 40, 42, 43, 44, 45, 51, 52, 53, 54, 56, 58, 59, 61, 66, 68, 69, 70, 73, 82) was used for HPV detection and typing following the manufacturer’s instructions. Each laboratory’s testing personnel completed competency assessment based on defined HPV plasmid samples prior initiating study testing (40). Results were analyzed using Seegene Viewer and formatted for REDCap. Assay validity was determined based on cycle threshold (Ct) values, and control samples included at each stage of the assay. For this study, samples were considered valid if at least one internal control or HPV type was detected. Samples were considered invalid if both internal controls were negative and no HPV was detected.

### Data entry, processing and statistical analysis

All data were entered into REDCap, and analyses conducted using SAS 9.4 (SAS Institute, Cary, NC) and Stata 18 (StataCorp LLC, College Station, TX). Continuous variables were summarized using the mean, standard deviation, median, minimum, and maximum values, while categorical variables were presented as counts and percentages, based on eligible participants with non-missing values. All statistical tests were two-sided with a significance level of 5%. HPV concordance between SCVS and FVU was evaluated across HPV categories, with emphasis on the 14 HR-HPV types (i.e. HPV 16, 18, 31, 33, 35, 39, 45, 51, 52, 56, 58, 59, 66, 68) and particularly HPV 16/18. Concordance was classified as full (identical HPV types or both samples negative), partial (at least one but not all HPV types matched), or full discordance (no common HPV types detected). Overall concordance was assessed using a 3×3 table (positive, negative, inadequate), while type-specific concordance was evaluated using positive and negative agreement, overall agreement, positive concordance, Cohen’s Kappa coefficient, and the McNemar test.

## Ethical considerations

Ethical approval was obtained from the relevant authorities in each participating country. In South Asia, approvals were secured from the Research Review and Ethical Review Committees of icddr,b (Bangladesh; protocol number: PR-23071); the Ethics Committee on Human Research of Aga Khan University (Pakistan; protocol number: 2023-9369-27297) and the National Bioethics Committee for Research (Pakistan; protocol number: NBCR-1011); and the Nepal Health Research Council (Nepal; protocol number: 619/2023). Additional ethical approvals were obtained from the Ethics Committee of the London School of Hygiene & Tropical Medicine (United Kingdom, LSHTM Ethics Ref: 29571) and the Institutional Review Board of the International Vaccine Institute (South Korea; IRB NUMBER: 2023-004) for each country-specific study. The study was conducted in accordance with the principles of the Declaration of Helsinki. Written informed consent was obtained from all participants aged 18 years and older. In Bangladesh, where legally married females under the age of 18 are recognized as adults under local law; written informed consent was obtained following local ethical and legal guidelines.

## Results

### HPV Prevalence by Sample Type

The overall HPV prevalence and concordance between SCVS and FVU samples were analyzed using data from 753 participants across three South Asian countries. No inadequate test results were reported from either sample type (Table 2). For HPV 16/18, detection rates were similar between sample types: 2.3% in SCVS and 2.4% in FVU. The detection rate for the 7 HR HPV vaccine types was 5.3% in SCVS samples and 5.0% in FVU samples. For the 9 HPV vaccine types and 14 HR-HPV types, the detection was slightly higher in SCVS samples (6.9% and 8.6%) compared to FVU samples (5.8% and7.2%). Prevalence varied by country but showed the same pattern. Nepal had the highest detection rates.

**Table 2.** HPV prevalence from SCVS vs FVU by country and overall.

| HPV Type | Bangladesh (N=250) |  | Pakistan (N=253) |  | Nepal (N=250) |  | Overall<sup>5</sup> (N=753) |  |
| --- | --- | --- | --- | --- | --- | --- | --- | --- |
|  | FVU<br>n<br>(%) | SCVS<br>n<br>(%) | FVU<br>n<br>(%) | SCVS<br>n<br>(%) | FVU<br>n<br>(%) | SCVS<br>n<br>(%) | FVU<br>n<br>(%) | SCVS<br>n<br>(%) |
| **HPV 16/18** | 3<br>(1.2%) | 2<br>(0.8%) | 4<br>(1.6%) | 4<br>(1.6%) | 11<br>(4.4%) | 11<br>(4.4%) | 18<br>(2.4%) | 17<br>(2.3%) |
| **7 HR HPV vaccine types<sup>1</sup>** | 6<br>(2.4%) | 8<br>(3.2%) | 9 (3.6%) | 8<br>(3.2%) | 23<br>(9.2%) | 24<br>(9.6%) | 38<br>(5.0%) | 40<br>(5.3%) |
| **9 HPV vaccine types<sup>2</sup>** | 7<br>(2.8%) | 12<br>(4.8%) | 12<br>(4.7%) | 13<br>(5.1%) | 25<br>(10.0%) | 27<br>(10.8%) | 44<br>(5.8%) | 52<br>(6.9%) |
| **14 HR-HPV types<sup>3</sup>** | 10<br>(4.0%) | 10<br>(4.0%) | 11<br>(4.3%) | 13<br>(5.1%) | 33<br>(13.2%) | 42<br>(16.8%) | 54<br>(7.2%) | 65<br>(8.6%) |
| **Any 28 HPV types<sup>4</sup>** | 15<br>(6.0%) | 20<br>(8.0%) | 22<br>(8.7%) | 26<br>(10.3%) | 56<br>(22.4%) | 69<br>(27.6%) | 93<br>(12.4%) | 115<br>(15.3%) |

<sup>1</sup> HPV 16, 18, 31, 33, 45, 52, 58 (7 HR HPV vaccine types). Note this category only includes the 7 HR HPV vaccine types covered by the nonavalent HPV vaccine and does not include HPV 6 and 11, which are considered low risk since they are not known to cause cancer.
<sup>2</sup> HPV 6, 11, 16, 18, 31, 33, 45, 52, 58 (9 HPV vaccine types). This category includes all 9 HPV vaccine types covered by the nonavalent HPV vaccine.
<sup>3</sup> HPV 16, 18, 31, 33, 35, 39, 45, 51, 52, 56, 58, 59, 66, 68 (14 HR-HPV types)
<sup>4</sup> HPV 6, 11, 16, 18, 26, 31, 33, 35, 39, 40, 42, 43, 44, 45, 51, 52, 53, 54, 56, 58, 59, 61, 66, 68, 69, 70, 73, 82 (any 28 HPV types). These are the 28 HPV types detectable by our Seegene Allplex28.
<sup>5</sup> Prevalence for combined data from all three South Asian countries.

### HPV Concordance by Sample Types

As the pattern of sample type concordance was similar for all countries aggregated results are shown (Table 3). Full concordance ranged from 99.3% (HPV 16/18) to 88.4% (28 HPV types) and full discordance ranged from 8.4% (38 HPV types) to 0.7% (HPV 16/18).

**Table 3.**
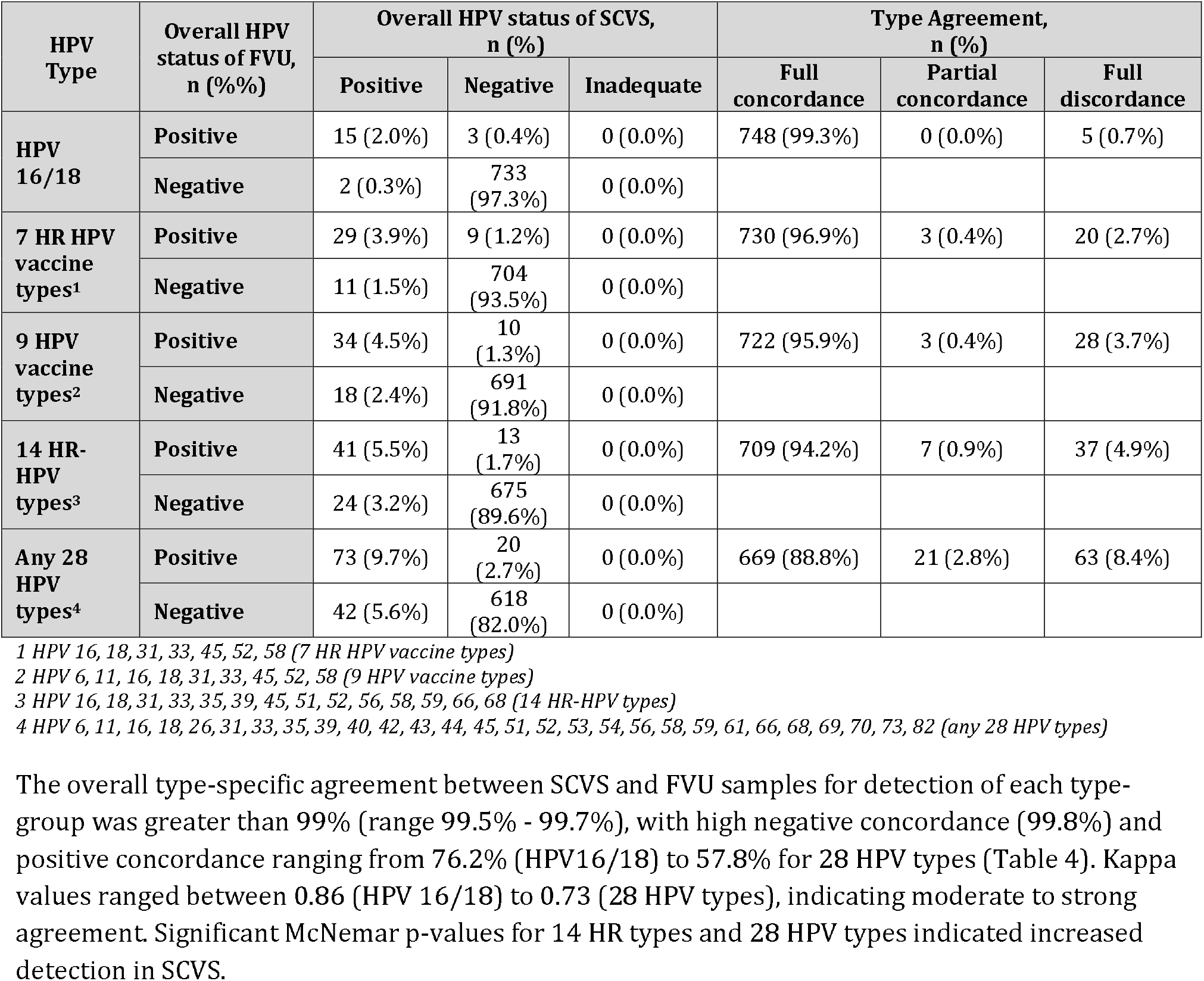
Overall HPV concordance between SCVS and FVU for all three South Asian countries.

| HPV Type | Overall HPV status of FVU, n (%) | Overall HPV status of SCVS, n (%) |  |  | Type Agreement, n (%) |  |  |
| --- | --- | --- | --- | --- | --- | --- | --- |
|  |  | Positive | Negative | Inadequate | Full concordance | Partial concordance | Full discordance |
| HPV 16/18 | Positive | 15 (2.0%) | 3 (0.4%) | 0 (0.0%) | 748 (99.3%) | 0 (0.0%) | 5 (0.7%) |
|  | Negative | 2 (0.3%) | 733 (97.3%) | 0 (0.0%) |  |  |  |
| 7 HR HPV vaccine types<sup>1</sup> | Positive | 29 (3.9%) | 9 (1.2%) | 0 (0.0%) | 730 (96.9%) | 3 (0.4%) | 20 (2.7%) |
|  | Negative | 11 (1.5%) | 704 (93.5%) | 0 (0.0%) |  |  |  |
| 9 HPV vaccine types<sup>2</sup> | Positive | 34 (4.5%) | 10 (1.3%) | 0 (0.0%) | 722 (95.9%) | 3 (0.4%) | 28 (3.7%) |
|  | Negative | 18 (2.4%) | 691 (91.8%) | 0 (0.0%) |  |  |  |
| 14 HR-HPV types<sup>3</sup> | Positive | 41 (5.5%) | 13 (1.7%) | 0 (0.0%) | 709 (94.2%) | 7 (0.9%) | 37 (4.9%) |
|  | Negative | 24 (3.2%) | 675 (89.6%) | 0 (0.0%) |  |  |  |
| Any 28 HPV types<sup>4</sup> | Positive | 73 (9.7%) | 20 (2.7%) | 0 (0.0%) | 669 (88.8%) | 21 (2.8%) | 63 (8.4%) |
|  | Negative | 42 (5.6%) | 618 (82.0%) | 0 (0.0%) |  |  |  |

<sup>1</sup> HPV 16, 18, 31, 33, 45, 52, 58 (7 HR HPV vaccine types)
<sup>2</sup> HPV 6, 11, 16, 18, 31, 33, 45, 52, 58 (9 HPV vaccine types)
<sup>3</sup> HPV 16, 18, 31, 33, 35, 39, 45, 51, 52, 56, 58, 59, 66, 68 (14 HR-HPV types)
<sup>4</sup> HPV 6, 11, 16, 18, 26, 31, 33, 35, 39, 40, 42, 43, 44, 45, 51, 52, 53, 54, 56, 58, 59, 61, 66, 68, 69, 70, 73, 82 (any 28 HPV types)

The overall type-specific agreement between SCVS and FVU samples for detection of each type-group was greater than 99% (range 99.5% −99.7%), with high negative concordance (99.8%) and positive concordance ranging from 76.2% (HPV16/18) to 57.8% for 28 HPV types (Table 4). Kappa values ranged between 0.86 (HPV 16/18) to 0.73 (28 HPV types), indicating moderate to strong agreement. Significant McNemar p-values for 14 HR types and 28 HPV types indicated increased detection in SCVS.

**Table 4.** Type-specific concordance between SCVS and FVU for all three South Asian countries.

| HPV 16/18 |  | SCVS(+) | SCVS(-) | Total | Proportion discrepant types, % | Positive Agreement, % (95% CI) | Negative Agreement, % (95% CI) | Overall agreement, % | Positive concordance, % | Kappa (95% CI) | Mc-Nemar p-value |
| --- | --- | --- | --- | --- | --- | --- | --- | --- | --- | --- | --- |
|  | FVU(+) | 16 | 3 | 19 | 0.33 | 88.9 (74.4 - 100) | 99.8 (99.6 - 100) | 99.7 | 76.2 | 0.86 (0.74 - 0.98) | 0.655 |
|  | FVU(-) | 2 | 1485 | 1487 |  |  |  |  |  |  |  |
|  | Total | 18 | 1488 | 1506 |  |  |  |  |  |  |  |
| 7 HR HPV vaccine types<sup>1</sup> |  | SCVS(+) | SCVS(-) | Total | Proportion discrepant types, % | Positive Agreement, % (95% CI) | Negative Agreement, % (95% CI) | Overall Agreement, % | Positive concordance, % | Kappa (95% CI) | Mc-Nemar p-value |
|  | FVU(+) | 39 | 11 | 50 | 0.44 | 76.5 (64.8 - 88.1) | 99.8 (99.7 - 99.9) | 99.6 | 62.9 | 0.77 (0.68 - 0.86) | 0.835 |
|  | FVU(-) | 12 | 5209 | 5221 |  |  |  |  |  |  |  |
|  | Total | 51 | 5220 | 5271 |  |  |  |  |  |  |  |
| 9 HPV vaccine types<sup>2</sup> |  | SCVS(+) | SCVS(-) | Total | Proportion discrepant types, % | Positive Agreement, % (95% CI) | Negative Agreement, % (95% CI) | Overall Agreement, % | Positive concordance, % | Kappa (95% CI) | Mc-Nemar p-value |
|  | FVU(+) | 48 | 13 | 61 | 0.49 | 70.6 (59.8 - 81.4) | 99.8 (99.7 - 99.9) | 99.5 | 59.3 | 0.74 (0.66 - 0.83) | 0.223 |
|  | FVU(-) | 20 | 6696 | 6716 |  |  |  |  |  |  |  |
|  | Total | 68 | 6709 | 6777 |  |  |  |  |  |  |  |
| 14 HR-HPV types<sup>3</sup> |  | SCVS(+) | SCVS(-) | Total | Proportion discrepant types, % | Positive Agreement, % (95% CI) | Negative Agreement, % (95% CI) | Overall Agreement, % | Positive concordance, % | Kappa (95% CI) | Mc-Nemar p-value |
|  | FVU(+) | 75 | 17 | 92 | 0.49 | 68.2 (59.5 - 76.9) | 99.8 (99.8 - 99.9) | 99.5 | 59.1 | 0.74 (0.67 - 0.81) | 0.013 |
|  | FVU(-) | 35 | 10415 | 10450 |  |  |  |  |  |  |  |
|  | Total | 110 | 10432 | 10542 |  |  |  |  |  |  |  |
| Any 28 HPV types<sup>4</sup> |  | SCVS(+) | SCVS(-) | Total | Proportion discrepant types, % | Positive Agreement, % (95% CI) | Negative Agreement, % (95% CI) | Overall Agreement, % | Positive concordance, % | Kappa (95% CI) | Mc-Nemar p-value |
|  | FVU(+) | 144 | 32 | 176 | 0.50 | 66.4 (60.1 - 72.6) | 99.8 (99.8 - 99.9) | 99.5 | 57.8 | 0.73 (0.68 - 0.78) | <0.001 |
|  | FVU(-) | 73 | 20835 | 20908 |  |  |  |  |  |  |  |
|  | Total | 217 | 20867 | 21084 |  |  |  |  |  |  |  |

[Note] 2x2 table is developed by summing the cases for each HPV type within the HPV-type group. Proportion discrepant types: Total number of discordant cases / Total. Positive agreement: Total number of positives detected by both SCVS and FVU / Total number of positives detected by SCVS. Negative agreement: Total number of negatives detected by both SCVS and FVU / Total number of negatives detected by SCVS. Overall agreement: Total number of positives and negatives by both FVU and SCVS / Total. Positive concordance: Total number of positives detected by both SCVS and FVU / Total number of positives detected by SCVS or FVU
<sup>1</sup> 7 HR HPV vaccine types: HPV 16, 18, 31, 33, 45, 52, 58; <sup>2</sup> 9 HPV vaccine types: HPV 6, 11, 16, 18, 31, 33, 45, 52, 58; <sup>3</sup> 14 HR-HPV types: HPV 16, 18, 31, 33, 35, 39, 45, 51, 52, 56, 58, 59, 66, 68; <sup>4</sup> Any 28 HPV types: HPV 6, 11, 16, 18, 26, 31, 33, 35, 39, 40, 42, 43, 44, 45, 51, 52, 53, 54, 56, 58, 59, 61, 66, 68, 69, 70, 73, 82

### Cycle threshold (Ct) value

Cycle threshold (Ct) values, which inversely reflect viral copy numbers, were compared between samples that yielded concordant and discordant HPV detection between SCVS and FVU samples. Data for Bangladesh, Pakistan, and Nepal, as well as all three countries combined are shown (Figures 1, Table 5). The 274 concordant cases had a mean Ct value of 28.8 (95% CI: 28.1–29.5, range 14.7-40.6) with similar results for either sample type. The 104 discordant cases had mean Ct values of 36.1 (95% CI: 35.1–37.0, range 18.5 −42.9); significantly higher than the Ct values of concordant samples (p < 0.001). Results for either sample type were similar. Results for each country analyzed separately show the same pattern (Supplementary Figure 1 A–C).

**Table 5.** Overall Ct value summary.

| Category | N | Mean (95% CI) | Median (Q1-Q3) | Min-Max |
| --- | --- | --- | --- | --- |
| **Concordant** | **274** | **28.8 (28.1-29.5)** | **28.5 (24.6-33.5)** | **14.7-40.6** |
| Concordant - SCVS | 137 | 27.9 (27.0-28.9) | 27.8 (23.8-31.6) | 15.3-39.5 |
| Concordant - FVU | 137 | 29.6 (28.6-30.6) | 30.2 (25.6-35.0) | 14.7-40.6 |
| **Discordant** | **104** | **36.1 (35.1-37.0)** | **37.6 (33.6-39.3)** | **18.5-42.9** |
| Discordant - SCVS | 72 | 36.2 (35.2-37.2) | 37.5 (33.5-39.3) | 23.7-42.6 |
| Discordant - FVU | 32 | 35.8 (33.6-38.0) | 37.8 (34.4-39.4) | 18.5-42.9 |

[Note] For the comparison two samples t-test was used.
- P-value for comparison Concordant vs. Discordant is $<0.001$ - P-value for comparison Concordant -SCVS vs Discordant - SCVS is $<0.001$ - P-value for comparison Concordant - FVU vs Discordant - FVU is $<0.001$

**Figure 1.**
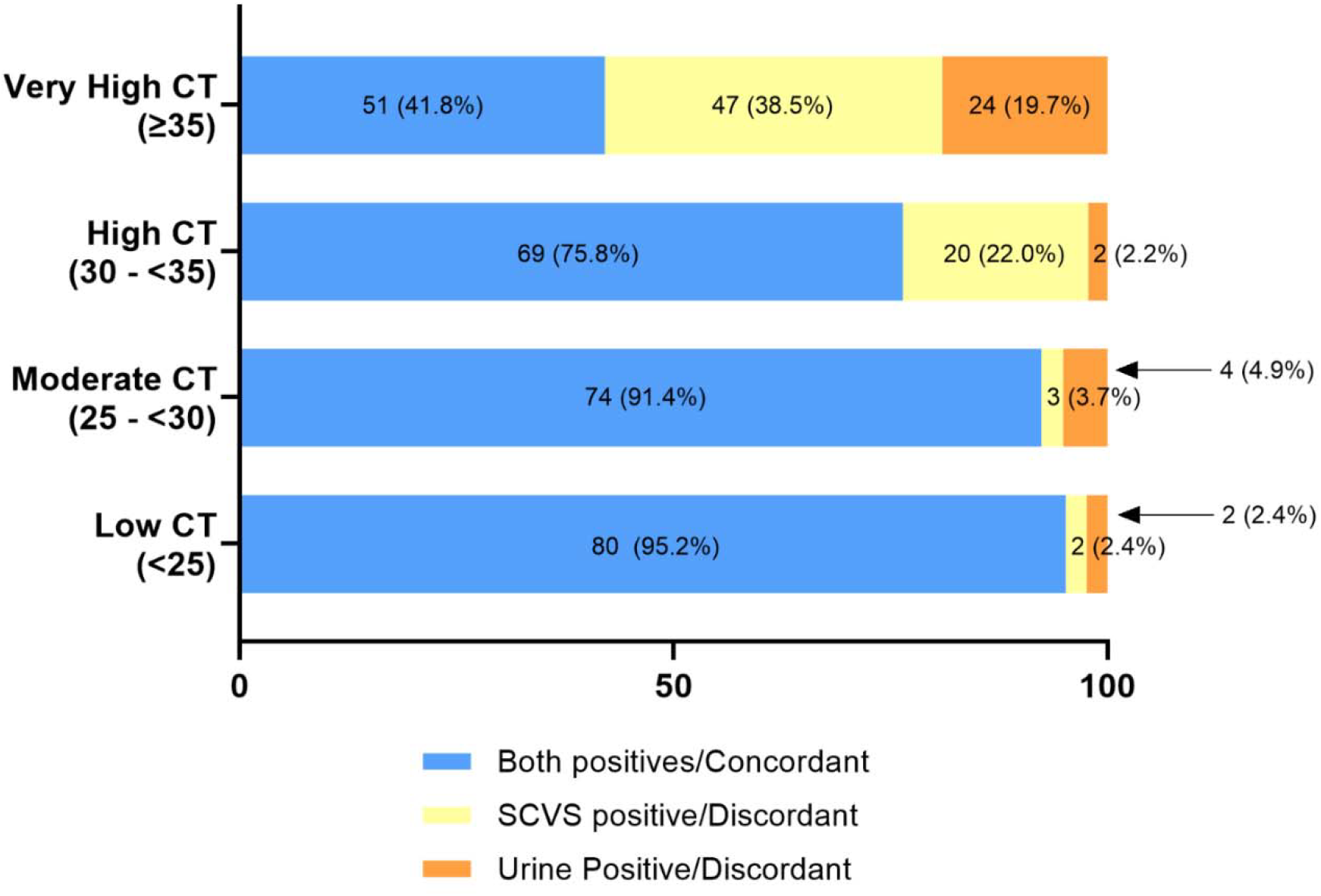
Rates of concordance and discordance between SCVS and FVU according to Ct value for the three South Asian countries.

## Discussion

In this study we evaluated the concordance between SCVS and FVU samples for HPV typing among sexually active girls and women from three South Asian countries: Bangladesh, Pakistan, and Nepal. Our findings provide strong evidence that both self-sampling methods provide comparable results for HPV detection, with SCVS demonstrating slightly higher sensitivity in detecting a broader range of HPV types, particularly non-vaccine HR and low risk types. These results demonstrate the use of self-sampling approaches, both FVU and SCV to assess changes in HPV genotype epidemiology and monitor vaccine impact (21, 23, 24).

The concordance between SCVS and FVU samples for HPV detection was similar across all study sites. The concordance for HPV 16/18 was very high, with overall agreement exceeding 99% across all study sites and Kappa values of 0.80 (95% CI: 0.75–0.85) in Bangladesh, 0.80 (95% CI: 0.76–0.84) in Pakistan, and 0.91 (95% CI: 0.88–0.94) in Nepal. For the combined data from the three countries, the concordance of HPV 16/18 detection between SCVS and FVU was also notably high, with a Kappa value of 0.86 (95% CI: 0.74– 0.98). These results align with previous studies highlight strong agreement between first void urine and cervical swab samples for detecting HPV 16/18 (21, 24, 41, 42).

We found substantial concordance between SCVS and FVU, with Kappa values of 0.77 (95% CI: 0.68–0.86) for 7 HR HPV vaccine types, 0.74 (95% CI: 0.66–0.83) for 9-valent types, and 0.74 (95% CI: 0.67–0.81) for 14 HR-HPV types. Positive agreement rates were 76.5% for 7 HR-HPV vaccine types, 70.6% for 9 HPV vaccine types, and 68.2% for 14 HR-HPV types, while negative agreement remained high (>99%). These findings are consistent with previous studies, which reported self-collected samples, including urine, reliably detect HR-HPV (21, 29, 36, 43, 44).

Notably, while the concordance of the HR-HPV vaccine types was high, we observed a slightly lower positive agreement for the broader HPV type groups, particularly those including non-vaccine and LR-HPV types. For any of the 28 HPV types, the Kappa value was 0.73 (95% CI: 0.68–0.78), with a positive agreement of 66.4% (95% CI: 60.1–72.6) and a negative agreement of 99.8% (95% CI: 99.8–99.9), aligning with previous studies demonstrating moderate-to-high agreement of FVU with cervical samples (24, 25, 35). Yang *et al*. reported Kappa < 0.6 for LR HPV types (44). On the other hand, other studies evaluating paired urine and cervical samples have reported slightly lower detection rates for LR-HPV types in vaginal and cervical samples than FVU collected with Colli-Pee device (29).

However, in contrast to our findings, some studies have reported that urine samples generally exhibit moderate to lower sensitivity than cervical and vaginal specimens for detecting HR-HPV types. A systematic review and meta-analysis noted that sensitivity was significantly higher when FVU samples were used compared to random or midstream urine (P = 0.004) (24). A study using the Cobas 4800 HPV test (a clinically validated PCR-based assay) using 5 – 50 mL first-stream urine reported a sensitivity of 68.6% for urine samples compared to 92.3% for cervical samples, while specificity remained high at 93.2% (45). Sample collection and processing has been shown to impact HPV detection in urine, attributable to several factors, including suboptimal timing of sample collection less than one hour after the last urination (21, 37); the use of random or midstream urine instead of FVU (24); absence or improper use of preservatives, inadequate storage conditions (46), inefficient sample concentration and DNA extraction methods, and variability in the sensitivity and specificity of the HPV detection assays employed (45, 47).

Our study also assessed Ct values, which are inversely related to viral load. concordant types had significantly lower Ct values than discordant types, suggesting that that discrepancies between SCVS and FVU detections are more likely when viral DNA concentrations are low, nearing the detection threshold of the assay. Although no previous studies have directly compared Ct values in cases of discordant HPV detection between self-collected urine and swab-based samples, Van Keer *et al*., reported good agreement in HPV viral load (DNA copies) between FVU samples and reference cervical samples (35). However, Payan *et al*. reported HPV viral loads to be approximately 50-fold higher in cervical samples than in urine (5.00 ± 1.73 vs. 3.77 ± 1.32 log/mL; p < 0.001)(48). Variations in reported copy numbers or Ct values can be attributed to differences in sample processing of both urine and swab (collection device, type and volume of media, extraction method). The vaginal swab, directly sampling the cervical vaginal pool, could have higher viral load, but using a large volume of sample collection media could dilute the sample. Urine collects cervical/vaginal cells indirectly, with highest concentrations in first void. Processing can further concentrate cells through pelleting, as used in our study. The HPV assay characteristics can also affect the agreement between sample methods, as sensitive amplification assays can mitigate differences in copy numbers. A recent study evaluating the Allplex HPV28 assay demonstrated that HR-HPV detection in FVU samples is accurate regardless of whether DNA is extracted manually or automatically and does not require urine pre-centrifugation (49).

## Conclusions

Both SCVS and FVU samples are reliable for HPV detection, with SCVS showing slightly higher sensitivity, especially as the number of HPV types increased. Despite minor differences, FVU demonstrated high concordance with SCVS, particularly for HPV 16/18, 7 HR-HPV, and 9-vaccine types. Higher viral loads (lower Ct values) were linked to stronger SCVS-FVU concordance.

## Strengths and Limitations

A major strength of this study is its large sample set from socioeconomically similar communities in three geographically distinct countries (Bangladesh, Pakistan, and Nepal), with harmonized sample collection and testing methods, enabling robust comparisons between SCVS and FVU for HPV detection. However, the results reflect HPV detection in young sexually active women aged 18–25 years, which may not be generalizable to other age groups. In addition, results with different sampling devices, media, processing, extraction and assays could differ.

## Data Availability

The data produced in the present study are available upon reasonable request.

## Supplementary Material

**Supplmentary Figure 1A.**
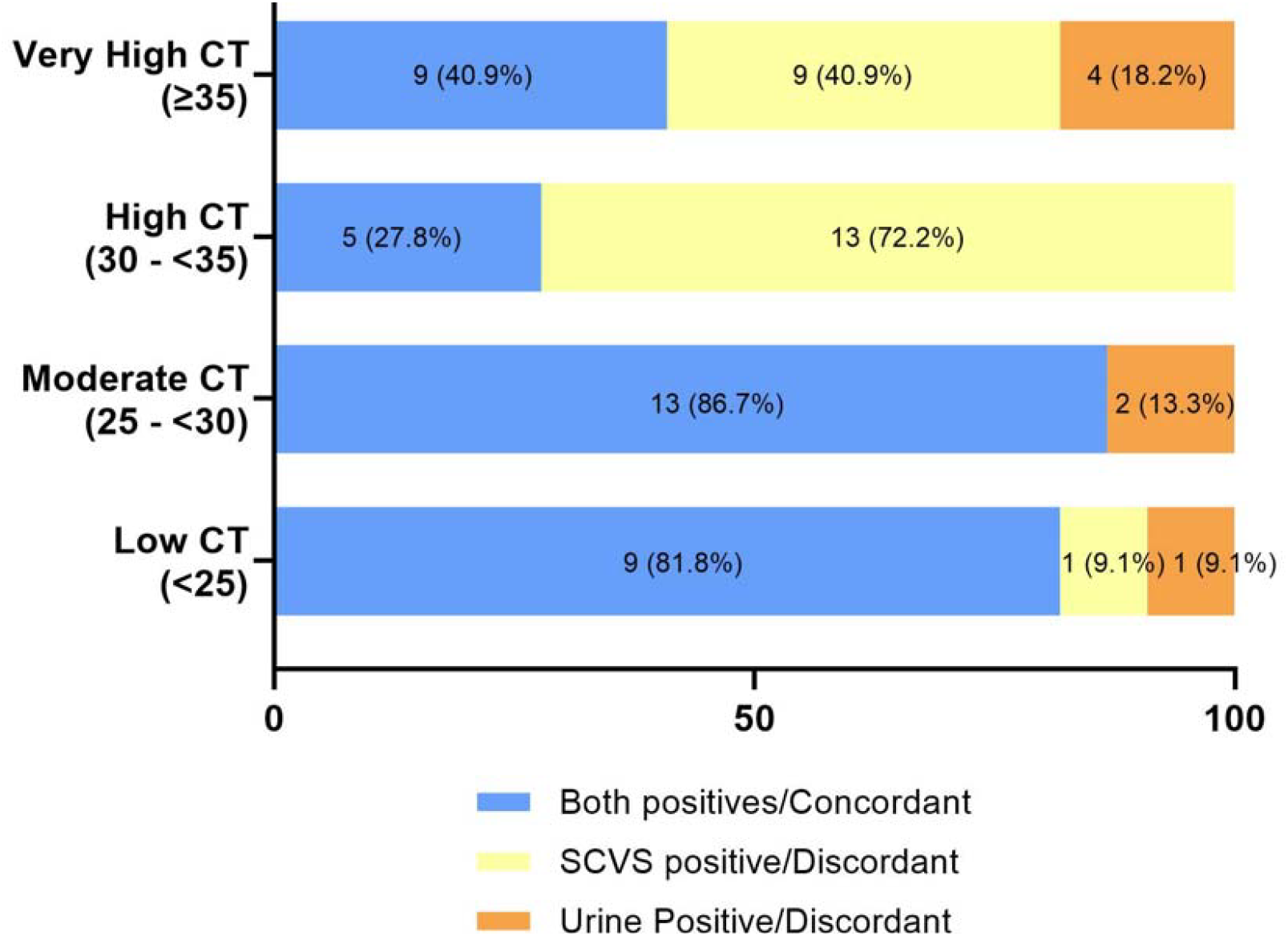
Rate of concordance and discordance between FVU and SCVS according to Ct value for Bangladesh.

**Supplmentary Figure 1B.**
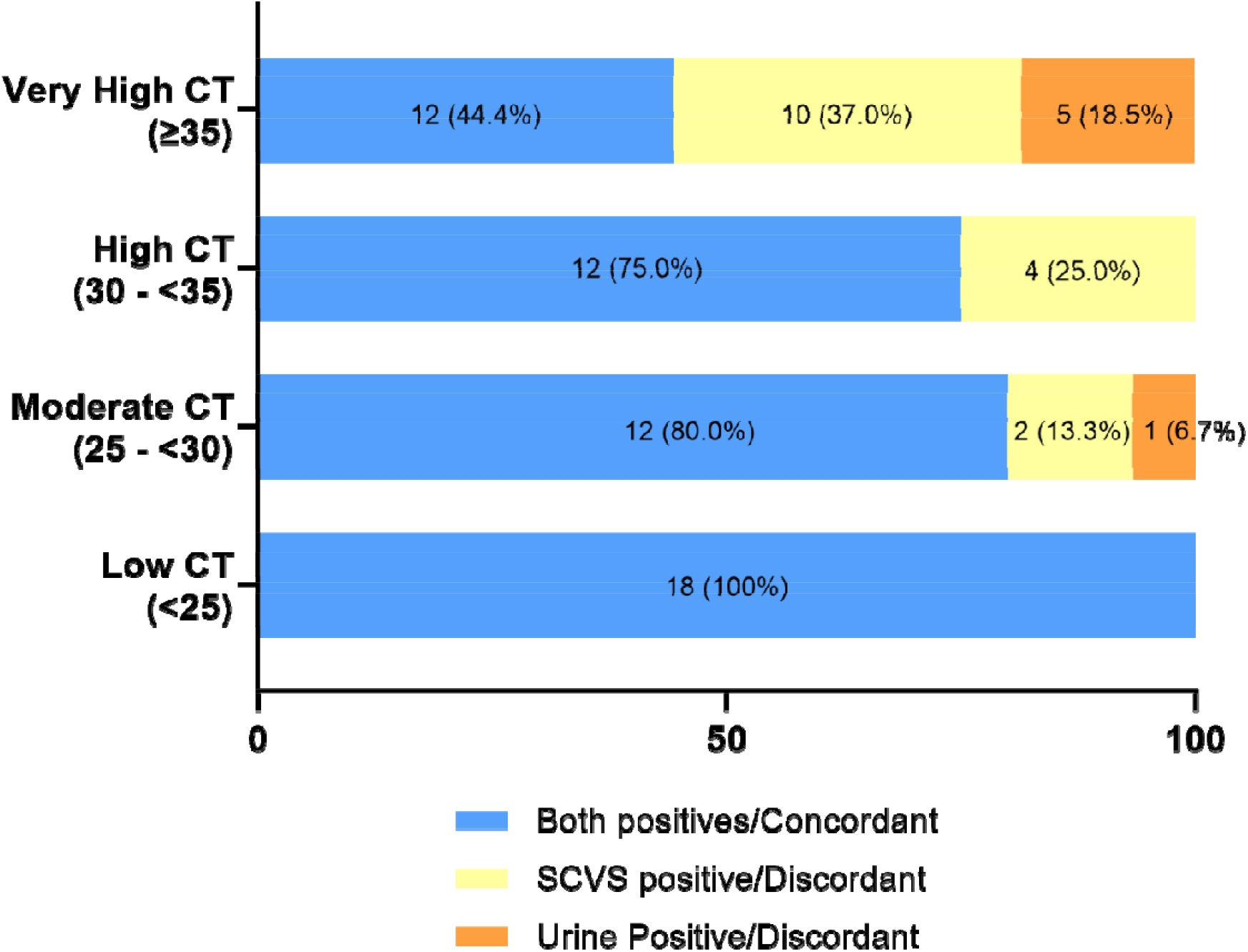
Rate of concordance and discordance between FVU and SCVS according to Ct value for Pakistan.

**Supplmentary Figure 1C.**
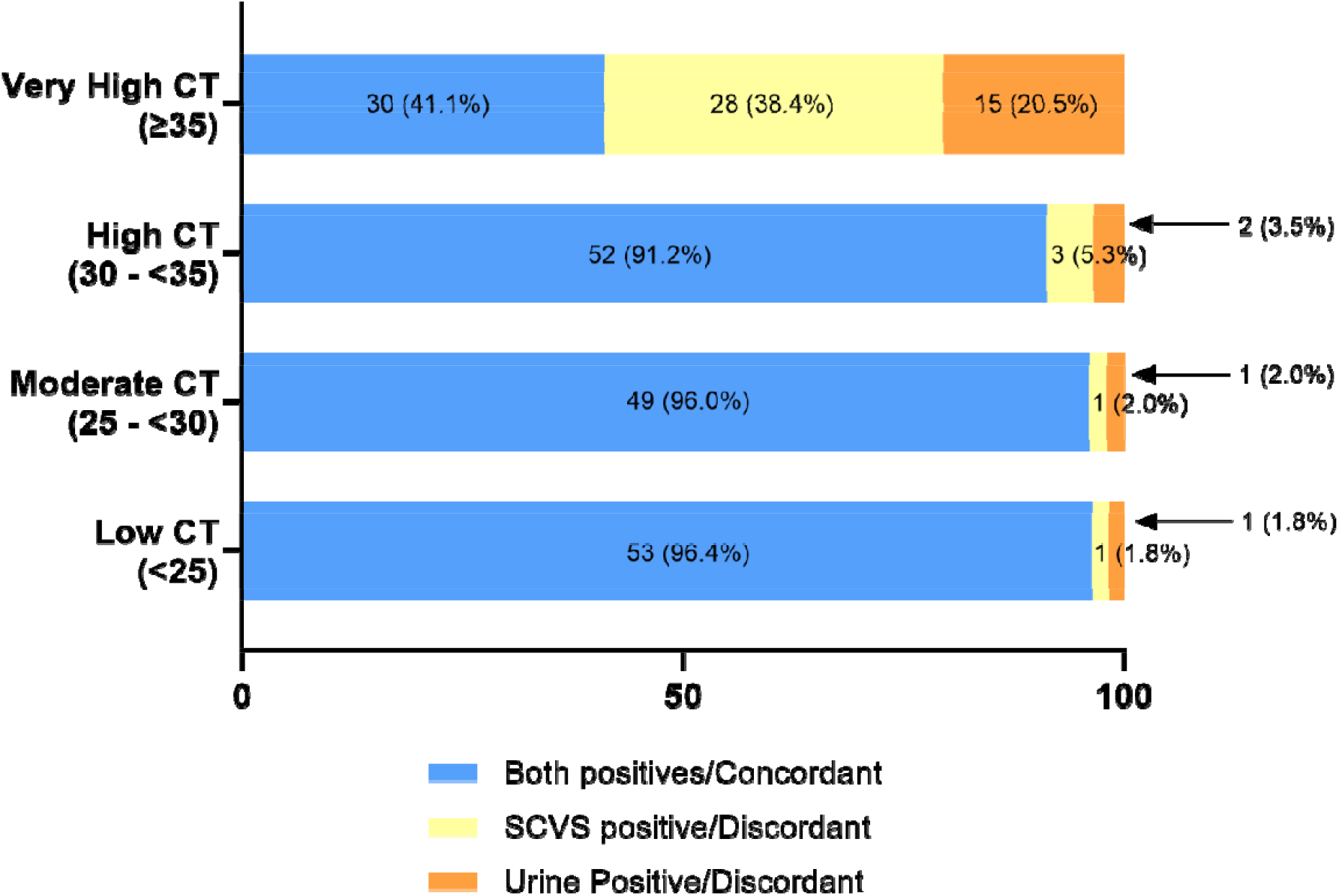
Rate of concordance and discordance between FVU and SCVS according to Ct value for Nepal.

